# Impact of the United Kingdom’s smokefree generation policy on tobacco-related equity in England: a simulation study

**DOI:** 10.1101/2025.06.13.25328563

**Authors:** Nathan P Davies, Rachael L Murray, Joanne R Morling, Manpreet Bains, Matthew Jones, Tessa Langley

## Abstract

**Objectives:** To explore how the United Kingdom’s smokefree generation policy may affect tobacco-induced inequalities in England by socio-economic status and sex.

**Design:** A yearly discrete-time, individual-level, state-transition microsimulation.

**Setting:** England from 2027 to 2075.

**Participants:** England general population born in or after 2009, including breakdown by Indices of Multiple Deprivation (IMD) quintile and sex.

**Interventions:** Four scenarios (baseline, pessimistic, central, optimistic) in which sale of tobacco is prohibited to those born in 2009 or later from 1 January 2027. Proportionate universalism sensitivity analyses in which intervention is 50% more effective in two most deprived quintiles and 50% less effective in two least deprived quintiles.

**Main outcome measures:** Smoking prevalence amongst 12-30-year-olds, equity of smoking prevalence by deprivation as measured by the slope index of inequality and relative index of inequality, and incremental quality adjusted life years (QALYs).

**Results:** The central scenario forecast smoking prevalence to be reduced to <5% in 12-30-year-olds by 2049, but not until 2055 for males and not until 2059 for those living in the most deprived quintile. Absolute socio-economic inequalities as measured by the slope index of inequality were reduced but not relative inequalities as measured by the relative index of inequality. Under a proportionate universalism scenario, <5% prevalence is achieved a year earlier (2048) and both absolute and relative inequalities by IMD quintile are non-significant by 2050. By 2075, 87,899 (85,293 – 90,791) discounted QALYs were gained in the central scenario compared to baseline.

**Conclusions:** The smokefree generation policy has potential to reduce absolute inequalities and achieve significant gains in quality and length of life. Achieving concurrent reductions in relative inequalities will likely require targeted interventions that lead to greater effectiveness in lower socio-economic areas and for males.

## BACKGROUND

Reaching the tobacco endgame involves moving beyond ongoing control of endemic tobacco use into a tobacco-free society.[1] Aiming for tobacco use rates below 5% is a realistic goal for nations with low rates of youth tobacco use.[2] Tobacco-free generation (TFG) or smoke-free generation (SFG) policies could play a role in achieving the tobacco endgame.[3] Policy formulations vary, but the core concept is to remove availability of tobacco - or in some cases, all nicotine products[4] - to those born after a certain cut-off birthdate, usually by banning retailers from selling products to this cohort.[5]

Recently, jurisdictions in the Philippines and several small towns in Massachusetts, United States (US) have introduced an SFG/TFG policy.[4,6,7] Although enforcement has begun in Brookline, US,[6] evaluation data is not yet available for these local policies, which will likely vary substantially from national-level policy. The best estimates of effect of national SFG policy on smoking rates and population health come from modelling studies.

Modelling studies based on SFG/TFG implementation in New Zealand and Singapore have projected that SFG will have some effect in the short-term on population prevalence, but other endgame tobacco control policies, such as denicotinisation, lead to greater early reductions.[8–10] Over the longer term, modelling from New Zealand, the Solomon Islands, Singapore, Canada, and England show that SFG/TFG is likely to have very large effects on smoking prevalence which will build over decades and achieve substantial gains in quality-adjusted life years (QALYs).[8–14] How rapidly SFG/TFG reduces population tobacco use prevalence compared to no implementation depends on the existing youth uptake rate for a nation, and – crucially – the assumption modellers make on the effect size of SFG/TFG. Some studies assume nobody born after the cut-off year will ever smoke[11,12,14] or large cumulative effects of the policy[13]; these simulations project relatively rapid reductions in smoking rate down towards zero. Others set a floor at which smoking continues at a very low proportion of existing smoking.[9] Some set the floor at the rate of illegal youth smoking at the time of modelling[8,10]; in these scenarios, population smoking rates reduce more slowly and level out above zero.

Despite projected gains from SFG/TFG policies, some researchers have raised concerns that SFG could result in increased health inequities, due to more challenging enforcement in areas with high tobacco prevalence and retail density, and potential lack of concurrent action to reduce smoking in older age groups, in whom smoking is concentrated in disadvantaged groups.[15] Some modelling studies consider absolute equity of outcome i.e. total differences in health gains between population groups. New Zealand-based studies projected greater health benefits for the Maori population than non-Maori, [9,12] especially amongst females,[9] and a Solomon Islands study found men gained more health in absolute terms.[11]

Determining differential impacts on population groups using an evidence-based estimation of effect is an important step for jurisdictions with aspirations of an equitable introduction of SFG/TFG. The United Kingdom (UK) could become the first nation-state to introduce an SFG policy, which will cover all tobacco products but not e-cigarettes.[16] The Tobacco and Vapes Bill, which includes a provision to introduce SFG from 2027 for those born in or after 2009, is progressing through UK Parliament.[16] Despite industry opposition[17] the policy attracts support amongst the young people it will affect, politicians and experts, and the public.[18–20] The UK government has modelled the introduction of SFG in England, developing five scenarios for year-on-year reductions in smoking initiation rates: a base case scenario, 10%, 30% (the central scenario), 60% and 90%. It estimated smoking prevalence in 2050 for 14-to 30-year-olds under these scenarios as 8.1%, 3.1%, 0%, 0% and 0% respectively, with respective social value gain of £0, £67 billion, £111 billion, £119 billion, and £121 billion, largely from improved productivity gains.[13]

Our study therefore aimed to explore how the SFG policy may have differential effects by socio-economic groups and sex in England, and to what extent it may mitigate or exacerbate tobacco-related health inequalities. We set out to use up-to-date estimates on what the effect of SFG may be on smoking prevalence by using the empirical literature on Tobacco 21 in the US and the theoretical literature on SFG.

## METHODS

### Model development and structure

We developed a discrete-time, individual-level, state-transition microsimulation model for the purposes of this study. The underlying code for the model uses R version 4.4.1 and is adapted from the Sick-Sicker health decision microsimulation model.[21] We report the model development against the University of Liverpool proposed quality assessment framework for tobacco models[22] (supplementary material A T1). Individuals can exist in one of four possible states in the model; non-smoker, current smoker, former smoker and the absorbing state of death (Figure 1). Each year, each alive individual may stochastically transition between states. E-cigarette use is not included in the model as the simultaneous introduction of new laws and powers relating to e-cigarettes in the Tobacco and Vapes Bill make it extremely complex to predict the ongoing relationship between tobacco and e-cigarette use.

**Figure 1:**
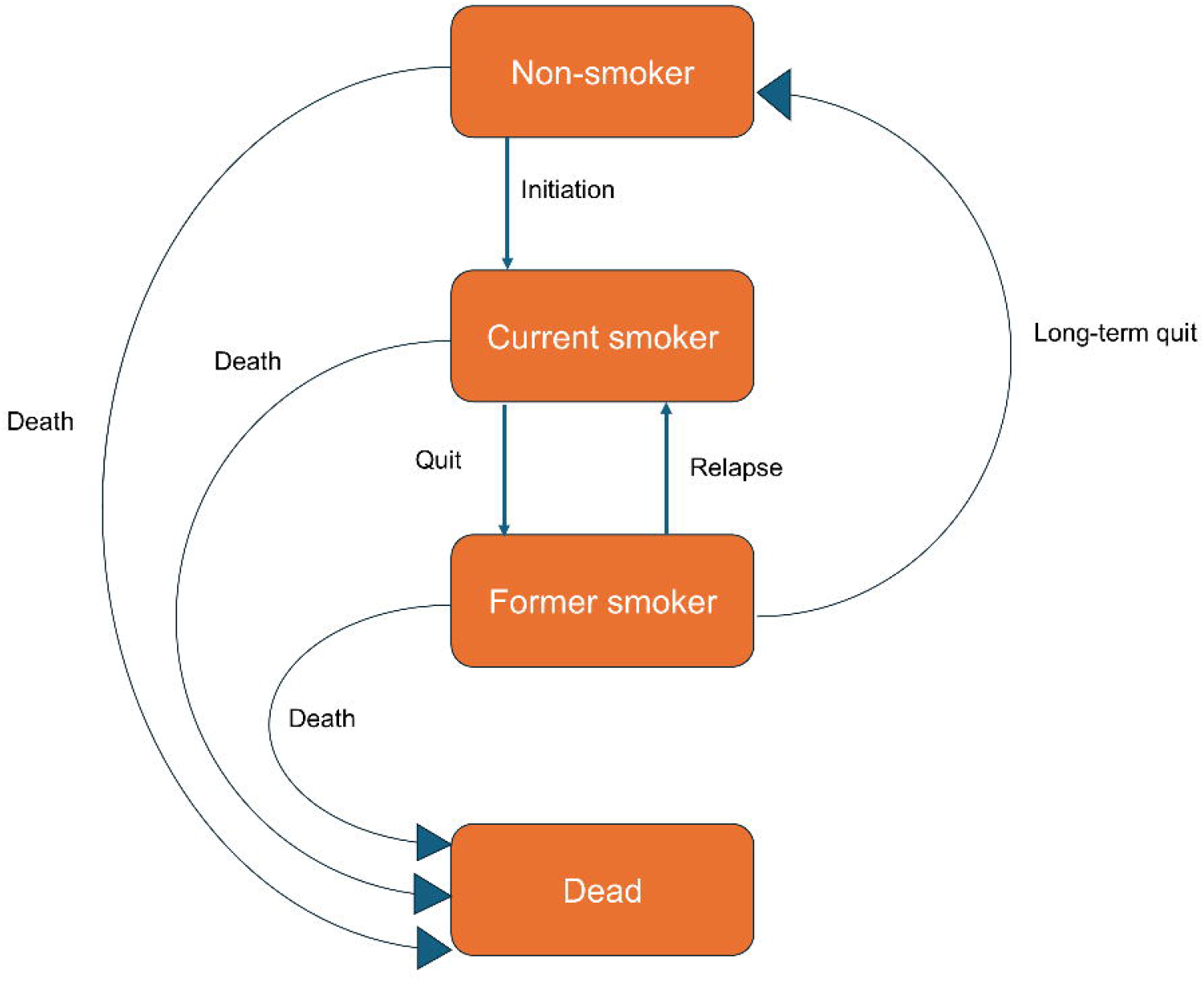
Schematic diagram of microsimulation model.

#### Starting population

The model uses ten representative 2% samples of individuals aged 11-30, as 30 is the last age non-smokers have any probability of moving to the smoking state. The sample uses single year of age and sex using figures from the Office for National Statistics (ONS) 2021-based interim national population projections for England.[23] Populating the model with a 1% sample projects smoking prevalences in 2050 that differ by fewer than 0.1 percentage points compared to a sample of 2% or 2.5% of the population by 2050. Ten samples of 2% enabled longer-term projections that vary by sex and index of multiple deprivation (IMD) status. Individuals were also assigned an IMD quintile, with the assumption individuals are equally distributed across quintiles by age and sex. New participants to the model are added aged 11 as non-smokers and subsequently progress through the model. A 2% sample of 700,000 new 11-year-olds are added each year - in line with 2021-based interim projections[23] – and evenly distributed across sex and IMD quintile. Point estimates are reported as the mean across the ten iterations.

#### Initial smoking probabilities

Initial smoking status by age and sex were taken from two sources. For 11-to 15-year-olds, data was taken from NHS England’s Smoking, Drinking and Drug Use among Young People in England[24] survey, and for adults aged 18+, the ONS Annual Population Survey was used.[25] Smoking statuses for 16- and 17-year-olds were imputed as the mean probabilities of 15- and 18-year-olds. Risk ratios for smoking by IMD quintile were calculated from the inequalities section of Office of Health Improvement and Disparities’ 2023 Smoking Profile[26] and applied as a weighted factor to the baseline probabilities. 11-year-olds were added into the model each year as non-smokers.

#### Smoking transition probabilities

The probabilities for moving between smoking states vary by age, sex and IMD quintile and are taken from the University of Sheffield’s estimations of initiation, quitting and relapse.[27] These are based on the Health Survey for England for initiation and quitting, and Hawkins et al for relapse rates up to 10 years after quitting (after which an individual becomes a non-smoker). In the base case scenario probabilities are estimated to remain constant at projected 2027 levels, based on the assumption no other major policies will be introduced that significantly reduce transition probabilities.

#### Mortality

Lifetable mortality probabilities were taken for each sex and year of age from ONS lifetables for the United Kingdom for the years 2017 – 2019 to avoid the effects of the COVID-19 pandemic.[28] As these data are not stratified by smoking status, all-cause mortality risk ratios calculated from 1987 – 2018 by Jeon et al were then used to modify the probability of death dependent on individual status as a current smoker, former smoker or non-smoker by sex and age bands from age 30 upwards.[29] The effects were assumed to be consistent across IMD quintiles.

#### Effect of smoke-free generation policy

Four scenarios for England have been modelled. In Scenario 1 (baseline), there is no introduction of SFG and smoking transition probabilities are held constant at 2027 levels.

In Scenario 2, the pessimistic scenario in which SFG is introduced, estimates of effect are calculated based on meta-analyses of recent studies of at least moderate quality from a systematic review of Tobacco 21[30] (details of methodology in Supplementary Material B). This results in an assumption of a fixed 14% reduction in initiation for those aged under 18 after the policy is introduced in 2027, and a fixed 23% reduction in initiation for those aged 18 or over. This assumes there is no ongoing cumulative effect of the policy on reducing initiation, based on two recent rigorous national studies of Tobacco 21 laws in the US which were examined multiple years’ worth of data on areas that had implemented Tobacco 21.[31,32] When examining the effect of Tobacco 21 at various timepoints since the new law was enacted, the size of effect appeared consistent over time for jurisdictions who introduced the law compared to jurisdictions that did not, and that effect size did not increase over a 2-year period compared to a single year period.

In Scenario 3, the central SFG scenario, a 5% annual cumulative decrease in smoking initiation was modelled on top of the reductions set out in Scenario 2. This cumulative trend effect is plausible for SFG beyond the fixed effects produced by Tobacco 21. SFG results in the age of sale continuing to rise year on year, continuing to affect individuals beyond the age of 21, which will in turn is likely to make it more difficult for younger adults to obtain cigarettes from older siblings, cousins or friends.[33]

Scenario 4, the optimistic scenario, used the assumptions made by the UK government in their base case modelling, which was based on expert elicitation. The UK government assumed a 30% *cumulative* reduction in initiation rates year on year.[13]

We also conducted sensitivity analyses in which enforcement, communication and support is assumed to strongly follow the principles of proportionate universalism, where SFG communication and enforcement is targeted at areas of greater deprivation, to explore whether this would affect reductions in total prevalence and inequalities between groups.[34] We assumed a 50% increase in the effect of SFG in the central and UK government scenarios in the two lowest IMD quintiles and a 50% decrease in effect of SFG in the two highest quintiles.

#### Model validation

Model predictions of smoking prevalence were comparable to pre-existing modelling using the same probabilities.[35] Long-term smoking rates were projected to level out in the no-intervention scenario for 12-30 year olds at approximately 11.1%, higher than the UK government’s no-intervention scenario of 8%.[13] One of the likely reasons for this is because 12- and 13-year-olds are included in our model and thus more individuals develop either current smoking or former smoking status as they go into their teenage years, with a subsequent inflationary effect on overall smoking rates. The UK government model also relied on estimates from US data to calculate smoking status probabilities for 14- and 15-year-olds, whereas our model uses updated University of Sheffield estimates from England data sources. Life expectancy in the model was validated against ONS data.[28]

#### Measuring equity

The chief purpose of the analysis was to estimate the equity impact of the policy in England. Four measures of inequality in smoking prevalence are used for IMD, with the slope index of inequality (SII) and the relative index of inequality (RII) serving as the primary measures for absolute and relative inequality respectively. The absolute concentration index and the relative concentration index are included as secondary measures to complement SII and RII.[36] Table 1 summarises these measures of inequality and their interpretation. The dates 2026, 2040, 2050 and 2075 were selected for comparison to explore how inequality in smoking rates may change over the short-, medium- and long-term. To estimate equity in smoking rates between males and females in the four scenarios, measures of prevalence difference were used.

**Table 1:** Measures of equity used in analysis.

| Measure of equity | Absolute or relative | Definition | Interpretation |
| --- | --- | --- | --- |
| Absolute concentration index (ACI) | Absolute | Calculates the distribution of a measure across population subgroups, in other words the degree to which a measure is concentrated among disadvantaged or advantaged subgroups. Subgroups are weighted by population size. | 0 = no inequality<br><0 = indicator concentrated in disadvantaged<br>>0 = indicator concentrated in advantaged<br>Larger values indicate greater inequality |
| Relative concentration index (RCI) | Relative | Calculated by dividing the ACI by the population mean to provide a relative measure of inequality concentration. | 0 = no inequality<br>-1 to <0 = indicator concentrated in disadvantaged<br>+1 to >0 = indicator concentrated in advantaged<br>Larger values indicate greater inequality |
| Slope index of inequality (SII) | Absolute | Using regression, this method estimates the **difference** in a measure between the most-advantaged and most-disadvantaged subgroup. Measures for all subgroups (not just most- and least-advantaged) are used in estimating the difference. | 0 = no inequality<br><0 = indicator concentrated in disadvantaged<br>>0 = indicator concentrated in advantaged<br>Larger values indicate greater inequality |
| Relative index of inequality (RII) | Relative | Using regression, this method estimates the **ratio** for a measure between the most-advantaged and most-disadvantaged subgroup. Measures for all subgroups (not just most- and least-advantaged) are used in estimating the difference. | 1 = no inequality<br><1 = indicator concentrated in disadvantaged<br>>1 = indicator concentrated in advantaged<br>Values are interpreted logarithmically<br>e.g. an RII of 0.1 has the same magnitude of inequality as an RII of 10. |

Adapted from Schlotheuber 2022[36]

#### Measures of morbidity and mortality

Quality-adjusted life years (QALYs) were calculated, using utility weights by from Vogel et al, which are stratified b smoking status, sex, and age for English populations.[37] QALYs for each SFG scenario were incrementally compared to baseline by IMD group, sex and for the population as a whole. Undiscounted analyses and analyses discounted from 2023 at 3.5% per year were conducted.

#### Public involvement

Three groups of young people aged 12–21 in England provided advice on prioritisation of populations and outcome measures before study commencement.

## RESULTS

Each smokefree generation scenario was projected to significantly reduce smoking prevalence amongst 12 – 30-year-olds (Figure 2). The central SFG scenario was projected to reduce smoking prevalence below 5% in this group by 2049, with the same milestone being achieved in 2037 in the UK government SFG scenario. In the pessimistic scenario, where the reduction in smoking rates is not cumulative, smoking rates level out at 9.8% compared to 11.3% in the baseline scenario. Smoking rates are at 1.4% in the central scenario in 2075.

**Figure 2:**
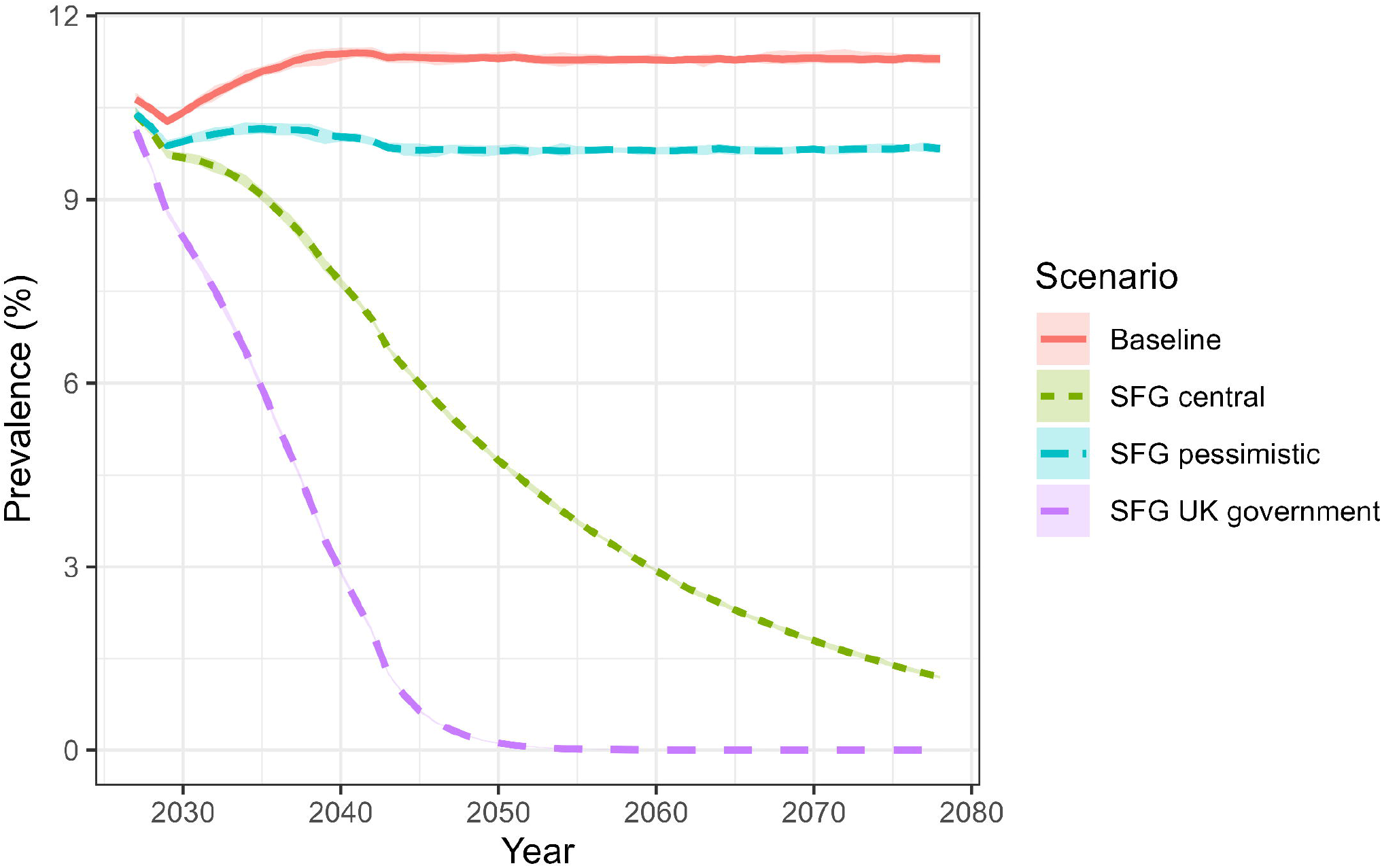
Projected current smoking prevalence amongst those aged 12-30 in England under baseline and three SFG scenarios with 95% first-order uncertainty intervals.

The smoking rate in the most deprived quintile takes longest to drop below 5% in the central scenario, doing so in 2059 (Figure 3). This compares to 2043 for those in the least deprived quintile. The smoking rate amongst males also takes longer to drop below 5%; in the central scenario, female prevalence goes below 5% in 2042, but male prevalence does not drop below 5% until 2055 (Supplementary material C F1).

**Figure 3:**
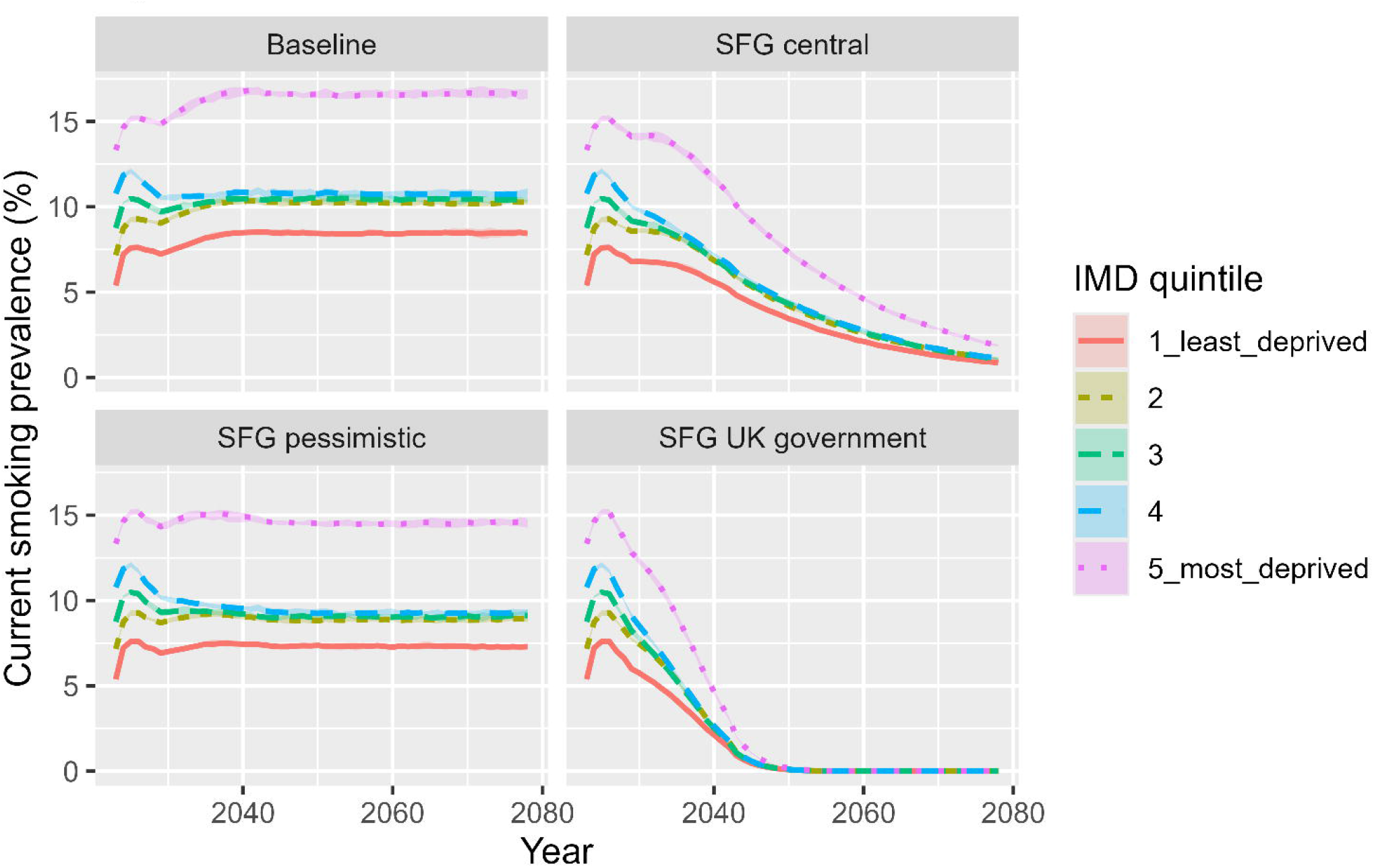
Projected current smoking prevalence amongst those aged 12 - 30 in England by IMD quintile under baseline and three SFG scenarios with 95% first-order uncertainty intervals.

Among individuals aged 12–30, the central SFG scenario reduces absolute inequalities in smoking prevalence (as measured by ACI and SII) by 2050 compared to 2026, and the UK government scenario reduces absolute inequalities by 2040 (Table 2). However, relative inequality measures (RII and RCI) showed minimal differences across both SFG scenarios. In the baseline scenario and in the pessimistic SFG scenario, there was no reduction in absolute or relative measures of inequality over time.

**Table 2:** IMD quintile equity measures for smoking prevalence for 12-30-year-olds by scenario and year.

| Strategy | Year | Absolute measures |  | Relative measures |  |
| --- | --- | --- | --- | --- | --- |
|  |  | SII | ACI | RII | RCI |
| Baseline | 2026 | -8.86 (-10.71, -7.02) | -1.41 (-1.80, -1.02) | 0.44 (0.38, 0.51) | -0.13 (-0.17, -0.09) |
| Baseline | 2040 | -8.60 (-14.00, -3.20) | -1.37 (-2.29, -0.44) | 0.47 (0.31, 0.70) | -0.12 (-0.20, -0.04) |
| Baseline | 2050 | -8.43 (-13.74, -3.13) | -1.34 (-2.25, -0.44) | 0.47 (0.32, 0.71) | -0.12 (-0.20, -0.04) |
| Baseline | 2075 | -8.45 (-13.83, -3.07) | -1.34 (-2.26, -0.42) | 0.47 (0.31, 0.71) | -0.12 (-0.20, -0.04) |
| SFG pessimistic | 2026 | -8.86 (-10.71, -7.02) | -1.41 (-1.80, -1.02) | 0.44 (0.38, 0.51) | -0.13 (-0.17, -0.09) |
| SFG pessimistic | 2040 | -7.76 (-12.67, -2.86) | -1.23 (-2.07, -0.40) | 0.46 (0.30, 0.70) | -0.12 (-0.21, -0.04) |
| SFG pessimistic | 2050 | -7.38 (-12.25, -2.50) | -1.17 (-2.00, -0.35) | 0.47 (0.31, 0.72) | -0.12 (-0.20, -0.04) |
| SFG pessimistic | 2075 | -7.56 (-12.45, -2.67) | -1.20 (-2.04, -0.36) | 0.46 (0.30, 0.70) | -0.12 (-0.21, -0.04) |
| SFG central | 2026 | -8.86 (-10.71, -7.02) | -1.41 (-1.80, -1.02) | 0.44 (0.38, 0.51) | -0.13 (-0.17, -0.09) |
| SFG central | 2040 | -6.27 (-10.36, -2.18) | -1.00 (-1.70, -0.29) | 0.44 (0.28, 0.69) | -0.13 (-0.22, -0.04) |
| SFG central | 2050 | -4.05** (-6.73, -1.37) | -0.64* (-1.10, -0.19) | 0.42 (0.27, 0.68) | -0.14 (-0.23, -0.04) |
| SFG central | 2075 | -1.24*** (-2.05, -0.44) | -0.20*** (-0.34, -0.06) | 0.41 (0.26, 0.66) | -0.14 (-0.24, -0.04) |
| SFG UK govt | 2026 | -8.86 (-10.71, -7.02) | -1.41 (-1.80, -1.02) | 0.44 (0.38, 0.51) | -0.13 (-0.17, -0.09) |
| SFG UK govt | 2040 | -2.59*** (-4.58, -0.61) | -0.41*** (-0.75, -0.07) | 0.41 (0.23, 0.72) | -0.14 (-0.26, -0.03) |
| SFG UK govt | 2050 | -0.10*** (-0.19, -0.02) | -0.02*** (-0.03, -0.00) | 0.41 (0.21, 0.77) | -0.14 (-0.27, -0.02) |
| SFG UK govt | 2075 | - | NA | NA | NA |

Parenthesis show 95% confidence intervals. \*Reduction in measure compared to 2026 significant at $p < 0.05$ \*\*Reduction significant at $p < 0.01$ \*\*\*Reduction significant at $p < 0.001$

In the proportionate universalism sensitivity analysis, 12-30-year-old population smoking prevalence was projected to drop below 5% a year earlier in both the main central scenario (2048) and the main UK government scenario (2036) (Supplementary Material C F2). In the proportionate universalism scenarios for the central scenario, absolute and relative inequality measures show no significant inequity by 2050; the measures then become unstable as prevalence approaches zero (Supplementary Material C T1).

All SFG scenarios lead to incremental QALYs gained compared to the baseline from 2050 onwards (Table 3). Incremental QALYs rise sharply in the 2060s and 2070s, given the long lead-time to most smoking-related death, and there are 87,899 (85,293 – 90,791) discounted incremental QALYs in the central scenario in 2075. Discounted QALYs gained in the pessimistic and central scenario in 2075 are 13.5% and 65.0% of those projected in the UK government scenario respectively. The most deprived quintile accounts for 28.0% of discounted QALYs gained by 2075 in the SFG central scenario and 33.0% in the SFG central proportionate universalism scenario (Supplementary material D T1). Males account for 66.4% of the QALYs gained in the central scenario (Supplementary material D T2).

**Table 3:** Incremental QALYS gained by scenario compared to baseline over time with first-order 95% uncertainty intervals.

| Year | Strategy | Incremental QALYs gained | Incremental QALYs gained (3.5% discount) |
| --- | --- | --- | --- |
| 2050 | SFG pessimistic | 13,859 (12,390 – 16,021) | 5,475 (4,894 – 6,329) |
| 2060 | SFG pessimistic | 31,219 (27,368 – 35,504) | 10,336 (9,169 – 11,571) |
| 2070 | SFG pessimistic | 51,756 (45,590 – 58,746) | 14,413 (13,003 – 16,034) |
| 2075 | SFG pessimistic | 74,865 (66,760 – 87,419) | 18,276 (16,745 – 20,809) |
| 2050 | SFG central | 53,995 (51,904 – 55,779) | 21,329 (20,503 – 22,034) |
| 2060 | SFG central | 136,144 (129,475 – 140,716) | 44,333 (42,235 – 45,772) |
| 2070 | SFG central | 247,753 (239,983 – 256,322) | 66,490 (64,243 – 68,642) |
| 2075 | SFG central | 375,838 (364,797 – 389,143) | 87,899 (85,293 – 90,791) |
| 2050 | SFG UK govt | 97,804 (94,121 – 100,524) | 38,634 (37,179 – 39,708) |
| 2060 | SFG UK govt | 226,282 (217,830 – 232,501) | 74,612 (71,822 – 76,587) |
| 2070 | SFG UK govt | 383,653 (370,801 – 392,819) | 105,853 (102,189 – 108,413) |
| 2075 | SFG UK govt | 559,133 (542,025 – 572,480) | 135,184 (130,809 – 138,364) |

## DISCUSSION

We find that, in our central SFG scenario, smoking prevalence within the 12-30 age group is projected to reach <5% in 2049. However, smoking prevalence in the most deprived quintile of postcodes does not reach <5% until 2059, and smoking prevalence in males does not do so until 2055. The central SFG scenario and UK government scenarios reduce measures of absolute inequality by IMD and by sex compared to baseline scenario, but measures of relative inequality stay constant.

As in other modelling studies on SFG, our study suggests that the policy is likely to have a positive effect on absolute inequality. Two simulations of the tobacco-free policy in Aotearoa/New Zealand project that SFG will lead to absolute reductions in smoking-related inequalities between those of Maori ethnicity and non-Maori ethnicity [9,12] with one study finding the per-capita health-adjusted life year (HALYs) gains are nine times higher for Maori females and 3.7 times higher for Maori males compared to non-Maoris.[9] Although we did not model by ethnicity, there are some parallels with our central scenario findings that SFG has the potential to reduce smoking-related absolute inequalities by IMD status. Modelling in the Solomon Islands found incremental lifetime HALY gains in males were around twice as high for males as females,[11] a finding closely reflected in QALY gains by sex in our study.

Our study differs from previous research by projecting differential effects on smoking initiation for SFG for groups experiencing health inequalities. Under the proportionate universalism central and UK government sensitivity analyses, in which enforcement effort, tailored communication and prevention efforts are presumed to be successfully targeted toward areas of higher deprivation, the gap between the most- and least-deprived quintiles closes almost completely by the time <5% prevalence is reached, and this milestone is reached one year earlier than the main scenarios. We assumed a 50% increased effect size in a proportionate universalist approach to implementing SFG, assuming targeted funding for communications, enforcement, and local work with young people can moderate the effects of the policy; this is illustrative rather than predictive. However, there is some evidence that it is plausible that raising the age of sale from 16 to 18 in England[38] and 18 to 21 in the US had greater effects on lower socio-economic groups.[39]

The strengths of our study design include England-specific smoking probabilities that vary by age, sex, and IMD status. The central scenario is based on empirical data from US T21 laws and accounts for theoretical cumulative effects of an SFG policy, and we also calculate pessimistic and optimistic scenarios and sensitivity analyses where the policy has differential effects across IMD groups. We use four measures of health inequity, including absolute and relative measures, and model scenarios in which proportionate universalism is applied, supporting understanding of the policy’s implications for equity.

One limitation is that simulating smoking rates decades into the future will always introduce significant uncertainties. It is difficult to know how social norms, retailer response and enforcement dynamics will develop with a novel intervention. SFG policy interactions with new e-cigarette laws and potential licensing approaches is not included in the model but are likely to be significant. There may also be some spatial and demographic clustering of tobacco use amongst specific location and groups as smoking prevalence trends down towards zero that we did not seek to model. We focused on policy effects on smoking rates, the largest source of uncertainty.

Our study has implications for how SFG is implemented in the UK and internationally. The Trading Standards workforce in England, who will be responsible for enforcing the age-of-sale policy, experienced a 39% real-terms reduction in funding last decade.[40] Under-18s in England continue to be sold tobacco by supermarkets and small shops even under current legislation[41] and funding and training for enforcement will be important in realising the potential of the policy to cumulatively and equitably reduce smoking prevalence as retail violations of age-of-sale laws are linked with higher youth smoking rates,[42,43] although people expect and desire for the law to be enforced.[19]

Further research may consider operational research into how enforcement should be implemented. It could also explore how other proposed policies - such as the introduction of tobacco licensing and new e-cigarette laws and policies - may also be used to complement SFG and target groups at greatest risk.

## Conclusion

SFG has the potential to substantially reduce smoking prevalence, gain hundreds of thousands of years of healthy life and reduce absolute tobacco-induced inequalities by deprivation and sex. With additional targeted action, it provides an opportunity to concurrently reduce relative inequalities. Our findings can inform both UK and global tobacco control strategies aiming for an equitable tobacco endgame.

## Data Availability

All data produced are contained in the manuscript and the source code of the model is available All data produced are available online at DOI:10.17605/OSF.IO/ANVQR

https://doi.org/10.17605/OSF.IO/ANVQR

## Ethical approval

Not required as this is a modelling study drawing on anonymised public data to create a synthetic population.

## Data availability statement

All data relevant to the study are included in the article or uploaded as supplemental information. All data used in the model are publicly available and their references provided in the manuscript. The model is available at OSF.

## Contributors

ND, RM, JM, TL and MJ were responsible for the study concept, and ND, RM, TL and MJ were responsible for the study design. ND was responsible for the model design and analysis. ND wrote the first draft of the manuscript. RM, JM, MB, TL and MJ critically revised the manuscript for important intellectual content.

## Transparency

ND and TL are the guarantors of this study. The corresponding author attests that all listed authors meet authorship criteria and that no others meeting the criteria have been omitted.

## Funding

This study is funded by the HEE/NIHR Integrated Clinical Academic Programme (grant NIHR302872). The views expressed are those of the authors and not necessarily those of the NIHR or the Department of Health and Social Care.

## Competing interests

None to declare.

## Dissemination to participants and related patient and public communities

The results of the study have been discussed with members of the public and civil society leaders who have provided ongoing advice and guidance on this study.

## Notes

### Competing Interest Statement

The authors have declared no competing interest.

### Author Declarations

The study used only openly available human data that are all provided in the OSF depository which holds the model code: DOI 10.17605/OSF.IO/ANVQR

