## Supplemental File A for "Impact of the United Kingdom’s smokefree generation policy on tobacco-related equity in England: a simulation study"

**Supplementary Material A, T1: Assessment against University of Liverpool tobacco modelling quality assessment**

| **Population:** model population data are representative of the population that the modelled policies will apply to. | The population has been derived from the Office for National Statistics, the government statistics agency for England. Smoking probabilities and QALY estimates are also England-specific. |
| --- | --- |
| **Policy effectiveness**: the policy effectiveness data were extracted from empirical evidence. | Policy effectiveness is based on a systematic review and meta-analysis of Tobacco 21 data, the closest existing policy. |
| **Smoking status**: the model captured the cumulative effect of smoking (smoking intensity, smoking history, quitting age, etc). | Model captures effect of former smoking for 10 years, but smoking intensity and pack-years are not built into the model. |
| **Smoking-related diseases**: the model estimated the effect on the majority of important smoking-related diseases (quantifying both morbidity and mortality). | The model estimates mortality based on a recent large-scale study of tobacco mortality. Morbidity is captured by use of QALY data stratified by age, sex and smoking status. |
| **Lag time**: the model explicitly captured the time lag between exposure and disease onset. | Mortality estimates are age-stratified, but do not account for age at which the individual started smoking. |
| **Transparency:** technical or non-technical documents available to provide model transparency. | Model described in detail in manuscript and R code for model made available on Open Science Framework. |
| **Uncertainty/sensitivity analysis** performed and reported. | First-order uncertainty from microsimulation draws reported.  Sensitivity analyses were performed:   - Three scenarios developed for potential effects of SFG on smoking - Proportionate universalism approaches also modelled in sensitivity analysis |
| **Validation**: the model was validated. | - Validated life expectancy of those in model to be consistent with ONS estimates - Validated smoking prevalence over long run in base case. When compared with the UK government model, it has higher prevalence for 14- to 30-year-olds, which is plausibly caused by different model assumptions (described in detail in main paper). |
| **Equity:** the model explored the equity impact of policies | This is the chief purpose of analysis and includes multiple technical measures of absolute and relative inequity by socio-economic status, and an exploration of a proportionate universalism approach to intervention implementation. |
