## Supplemental File B for "Impact of the United Kingdom’s smokefree generation policy on tobacco-related equity in England: a simulation study"

**Supplementary Material B: Estimation of Tobacco 21 effect**

**Methods**

We identified national-level effect estimates of Tobacco 21 (T21) laws from studies rated at “moderate” risk of bias within a recent systematic review of Tobacco 21.^[1]^ To be included in this meta-analysis, these estimates had to (1) be derived from nationally representative survey data extending to at least 2019, and (2) distinguish between participants aged ≥18 years and those under 18 years.

We extracted or calculated risk ratios (RR) for current smoking rate from these papers, using multiple model estimates where papers had used more than one technique. We performed the meta-analysis in R using the *metafor* package and used a multilevel modelling approach (via rma.mv) that included a random intercept for the variable “Dataset” to account for non-independence when authors analysed the same underlying survey data in multiple estimates. The restricted maximum likelihood (REML) method was used to estimate between-study variance. Heterogeneity was assessed with the Q statistic.

We ran separate meta-analyses for (a) ≥18 years, (b) a combined group of 8th and 10th graders (under 18). Effect sizes are presented as pooled RRs and 95% confidence intervals (CIs), with tests of heterogeneity reported by the Q statistic p-value.

**Results**

*≥18 years*
Six estimates from three datasets met our criteria for participants aged 18–20 years or older. The pooled effect indicated a statistically significant reduction in smoking following T21 implementation (ln(RR) = –0.26, *p* = 0.0145). Transformed back to the risk ratio scale, the combined point estimate was RR = 0.77 (95% CI: 0.62–0.95). The Q statistic suggested moderate heterogeneity (*p* = 0.075), indicating some variability across datasets.

*Under 18 (8th and 10th graders combined)*
Five effect estimates across two datasets were analysed jointly for 8th and 10th graders. The pooled effect was ln(RR) = –0.15 (*p* = 0.0307), corresponding to a RR of 0.86 (95% CI: 0.75–0.99). Heterogeneity in this analysis was low, with *p* = 0.663 for the Q statistic.

1. Nathan Davies, Ilze Bogdanovica, Shaun McGill, Rachael L Murray, What is the Relationship Between Raising the Minimum Legal Sales Age of Tobacco Above 20 and Cigarette Smoking? A Systematic Review, *Nicotine & Tobacco Research*, Volume 27, Issue 3, March 2025, Pages 369–377,
