## Supplemental File C for "Impact of the United Kingdom’s smokefree generation policy on tobacco-related equity in England: a simulation study"

**Supplementary Material C Figure 1: Projected current smoking prevalence amongst those aged 12 - 30 in England by sex under baseline and three SFG scenarios with 95% first-order uncertainty intervals**

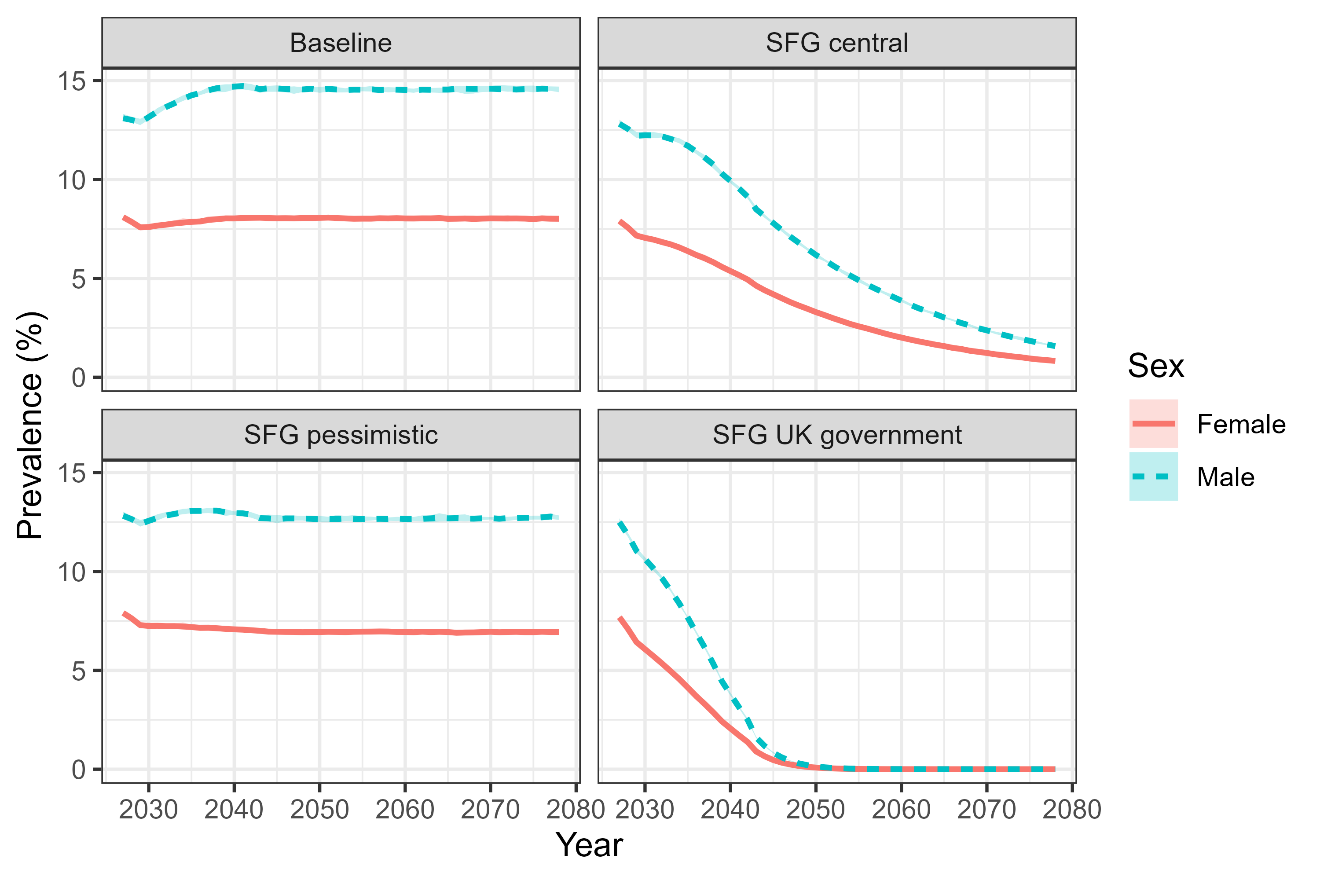

**Supplementary Material C Figure 2: Projected current smoking prevalence amongst those aged 12 - 30 in England by IMD under baseline and SFG proportionate universalism scenarios with 95% first-order uncertainty intervals**

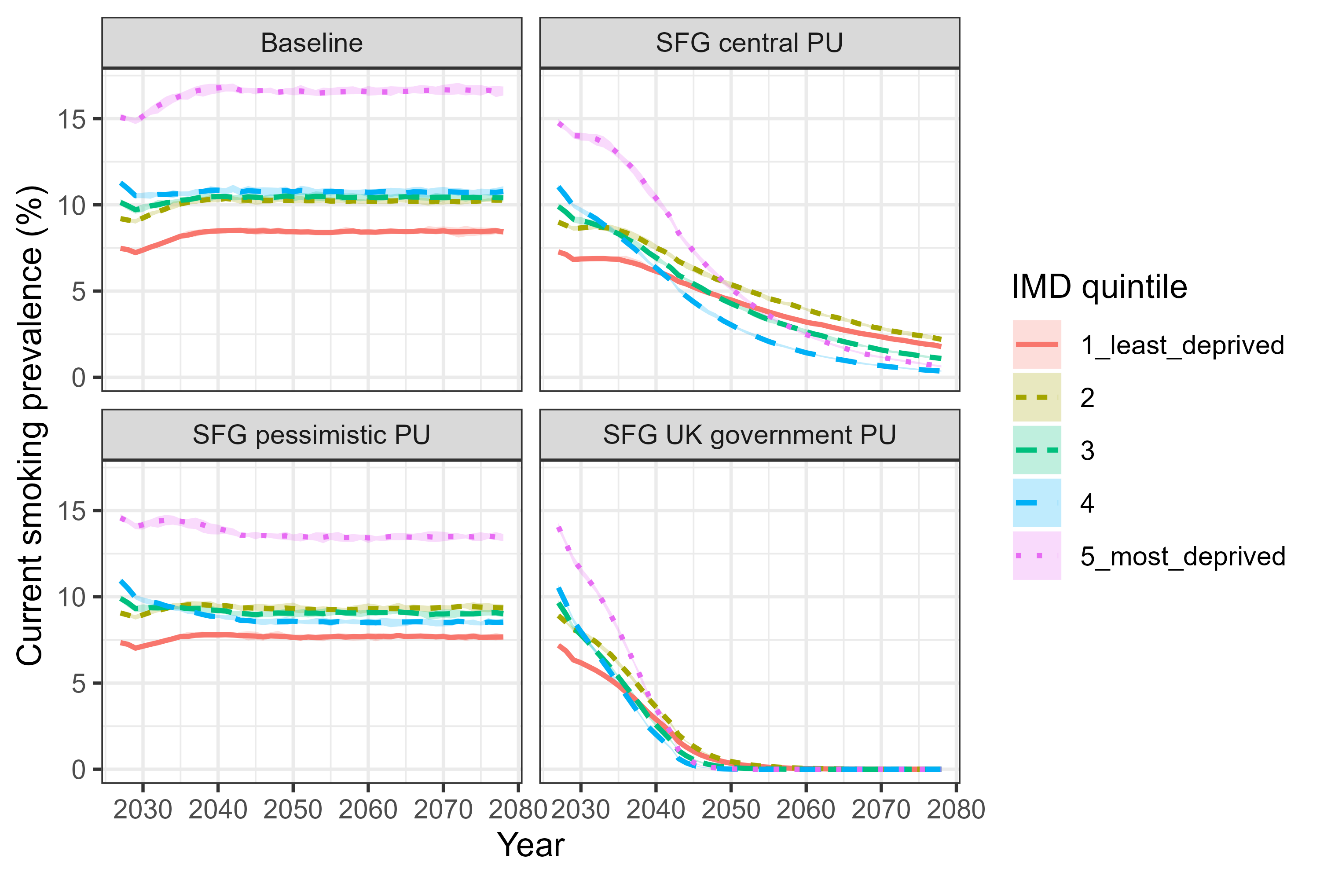

**Supplementary C Material T1: IMD quintile equity measures for smoking prevalence for 12 – 30-year-olds by scenario and year, proportionate universalism sensitivity analysis**

| **Strategy** | **Year** | **Absolute measures** | | **Relative measures** | |
| --- | --- | --- | --- | --- | --- |
|  |  | **SII** | **ACI** | **RII** | **RCI** |
| Baseline | 2026 | -8.86 (-10.71, -7.02) | -1.41 (-1.80, -1.02) | 0.44 (0.38, 0.51) | -0.13 (-0.17, -0.09) |
| Baseline | 2040 | -8.60 (-14.00, -3.20) | -1.37 (-2.29, -0.44) | 0.47 (0.31, 0.70) | -0.12 (-0.20, -0.04) |
| Baseline | 2050 | -8.43 (-13.74, -3.13) | -1.34 (-2.25, -0.44) | 0.47 (0.32, 0.71) | -0.12 (-0.20, -0.04) |
| Baseline | 2075 | -8.45 (-13.83, -3.07) | -1.34 (-2.26, -0.42) | 0.47 (0.31, 0.71) | -0.12 (-0.20, -0.04) |
| SFG pessimistic PU | 2026 | -8.86 (-10.71, -7.02) | -1.41 (-1.80, -1.02) | 0.44 (0.38, 0.51) | -0.13 (-0.17, -0.09) |
| SFG pessimistic PU | 2040 | -5.85 (-10.95, -0.75) | -0.93 (-1.79, -0.08) | 0.55 (0.35, 0.87) | -0.09 (-0.18, -0.01) |
| SFG pessimistic PU | 2050 | -5.44 (-10.35, -0.52) | -0.87 (-1.69, -0.05) | 0.57 (0.36, 0.89) | -0.09 (-0.18, -0.00) |
| SFG pessimistic PU | 2075 | -5.41 (-10.56, -0.26) | -0.86 (-1.73, 0.00) | 0.57 (0.35, 0.91) | -0.09 (-0.18, 0.00) |
| SFG central PU | 2026 | -8.86 (-10.71, -7.02) | -1.41 (-1.80, -1.02) | 0.44 (0.38, 0.51) | -0.13 (-0.17, -0.09) |
| SFG central PU | 2040 | -3.65* (-8.03, 0.72) | -0.58* (-1.31, 0.14) | 0.61 (0.36, 1.04) | -0.08 (-0.18, 0.02) |
| SFG central PU | 2050 | 0.53*** (-2.47, 3.54) | 0.08*** (-0.43, 0.60) | 1.13 (0.57, 2.24) | 0.02* (-0.10, 0.13) |
| SFG central PU | 2075 | 2.25*** (0.53, 3.98) | 0.34*** (0.09, 0.60) | 5.09*** (1.69, 15.38) | 0.25*** (0.06, 0.43) |
| SFG UK govt PU | 2026 | -8.86 (-10.71, -7.02) | -1.41 (-1.80, -1.02) | 0.44 (0.38, 0.51) | -0.13 (-0.17, -0.09) |
| SFG UK govt PU | 2040 | 0.16*** (-2.04, 2.37) | 0.03*** (-0.36, 0.41) | 1.06 (0.49, 2.27) | 0.01* (-0.12, 0.14) |
| SFG UK govt PU | 2050 | NA | NA | NA | NA |
| SFG UK govt PU | 2075 | NA | NA | NA | NA |

*Parenthesis show 95% uncertainty intervals. *Reduction in measure compared to 2026 significant at p < 0.05	**Reduction significant at p < 0.01	***Reduction significant at p < 0.001*
