## Supplemental File D for "Impact of the United Kingdom’s smokefree generation policy on tobacco-related equity in England: a simulation study"

**Supplementary Material D T1: Incremental QALYs gained by scenario by IMD quintile compared to baseline in 2075 (parenthesis show first-order 95% uncertainty intervals)**

| Year | Strategy | IMD quintile | Incremental QALYs gained | Incremental QALYs gained (3.5% disc) |
| --- | --- | --- | --- | --- |
| 2075 | SFG pessimistic | 1 | 11,912 (-6,842 – 30,063) | 3,071 (-669 – 6,370) |
| 2075 | SFG pessimistic | 2 | 12,811 (3,925 – 23,331) | 3,179 (1,454 – 5,603) |
| 2075 | SFG pessimistic | 3 | 11,370 (-4,157 – 27,777) | 2,720 (-637 – 6,156) |
| 2075 | SFG pessimistic | 4 | 13,072 (5,954 – 20,537) | 3,261 (2,112 – 5,059) |
| 2075 | SFG pessimistic | 5 | 25,701 (14,434 – 41,066) | 6,045 (4,055 – 9,452) |
| 2075 | SFG pessimistic PU | 1 | 6,090 (-16,735 – 26,621) | 1,609 (-3,050 – 5,438) |
| 2075 | SFG pessimistic PU | 2 | 7,486 (-4,707 – 16,563) | 2,074 (-664 – 3,980) |
| 2075 | SFG pessimistic PU | 3 | 12,289 (-922 – 27,212) | 2,885 ( 132 – 6,181) |
| 2075 | SFG pessimistic PU | 4 | 25,269 (10,507 – 37,260) | 6,085 (3,274 – 8,775) |
| 2075 | SFG pessimistic PU | 5 | 40,531 (27,889 – 49,071) | 9,556 (6,778 – 11,345) |
| 2075 | SFG central | 1 | 64,775 (42,412 – 81,219) | 15,166 (10,700 – 18,396) |
| 2075 | SFG central | 2 | 66,114 (53,022 – 77,576) | 15,532 (13,049 – 17,791) |
| 2075 | SFG central | 3 | 65,564 (51,524 – 83,391) | 15,338 (12,558 – 19,070) |
| 2075 | SFG central | 4 | 72,702 (64,507 – 82,805) | 17,227 (15,445 – 19,046) |
| 2075 | SFG central | 5 | 106,682 (96,285 – 114,590) | 24,635 (22,587 – 26,413) |
| 2075 | SFG central PU | 1 | 51,103 (26,867 – 70,960) | 11,961 (7,162 – 16,356) |
| 2075 | SFG central PU | 2 | 52,547 (32,920 – 72,283) | 12,448 (8,393 – 16,490) |
| 2075 | SFG central PU | 3 | 66,489 (34,780 – 94,901) | 15,529 (9,303 – 21,303) |
| 2075 | SFG central PU | 4 | 89,171 (76,677 – 96,577) | 21,064 (18,459 – 22,754) |
| 2075 | SFG central PU | 5 | 128,817 (119,503 – 139,328) | 29,935 (27,938 – 32,205) |
| 2075 | SFG UK government | 1 | 97,239 (69,606 – 119,058) | 23,430 (18,366 – 28,278) |
| 2075 | SFG UK government | 2 | 98,920 (80,116 – 129,817) | 24,056 (20,407 – 29,774) |
| 2075 | SFG UK government | 3 | 100,884 (87,951 – 118,679) | 24,489 (22,432 – 28,061) |
| 2075 | SFG UK government | 4 | 99,912 (61,919 – 117,404) | 24,459 (17,247 – 27,638) |
| 2075 | SFG UK government | 5 | 162,177 (143,215 – 182,301) | 38,751 (34,230 – 42,553) |
| 2075 | SFG UK government PU | 1 | 92,082 (61,368 – 121,548) | 22,138 (16,100 – 28,140) |
| 2075 | SFG UK government PU | 2 | 87,417 (67,943 – 102,923) | 21,278 (17,623 – 24,508) |
| 2075 | SFG UK government PU | 3 | 97,951 (74,615 – 126,840) | 23,833 (19,092 – 29,920) |
| 2075 | SFG UK government PU | 4 | 105,451 (79,165 – 120,681) | 25,895 (21,143 – 29,357) |
| 2075 | SFG UK government PU | 5 | 172,372 (161,314 – 195,728) | 41,171 (38,655 – 46,499) |

**Supplementary Material D T2: Incremental QALYs gained by scenario by sex compared to baseline in 2075 (parenthesis show first-order 95% uncertainty intervals)**

| Year | Strategy | Sex | Incremental QALYs gained | Incremental QALYs gained (3% disc) |
| --- | --- | --- | --- | --- |
| 2075 | SFG pessimistic | Female | 24,152 (13,422 – 38,399) | 6,317 (4,083 – 9,299) |
| 2075 | SFG pessimistic | Male | 50,713 (34,281 – 64,327) | 11,958 (8,349 – 15,130) |
| 2075 | SFG central | Female | 123,122 (106,200 – 143,289) | 29,508 (26,384 – 33,036) |
| 2075 | SFG central | Male | 252,716 (229,984 – 276,700) | 58,391 (54,337 – 63,214) |
| 2075 | SFG UK government | Female | 176,105 (158,936 – 193,401) | 43,792 (40,434 – 47,637) |
| 2075 | SFG UK government | Male | 383,027 (361,719 – 408,278) | 91,392 (86,571 – 96,826) |
